# Laboratory findings in coronavirus disease 2019 (COVID-19) patients: a comprehensive systematic review and meta-analysis

**DOI:** 10.1101/2020.06.07.20124602

**Authors:** Mohammad Karimian, Amirreza Jamshidbeigi, Gholamreza Badfar, Milad Azami

## Abstract

**Background:** In early December 2019, the first patient with COVID-19 pneumonia was found in Wuhan, Hubei Province, China. Recent studies have suggested the role of primary laboratory tests in addition to clinical symptoms for suspected patients, which play a significant role in the diagnosis of COVID-19. Therefore, the present study was conducted to evaluate laboratory findings in COVID-19 patients.

**Data Sources:** PubMed/Medline, Scopus, EMBASE, Web of Science (ISI), Cochrane Library, Ovid, Science Direct, CINAHL and EBSCO.

**Study Selection:** Cross-sectional of adverse outcomes stratified by the status of ICLs were selected.

**Data Extraction:** The prevalence of available variables for laboratory tests were extracted.

**Results:** Finally, 52 studies involving 5490 patients with COVID-19 entered the meta-analysis process. The prevalence of leukopenia, lymphopenia, elevated c-reactive protein (CRP), elevated erythrocyte sedimentation rate (ESR), elevated serum amyloid A, elevated ferritin was estimated to be 20.9% (95%CI: 17.9-24.3), 51.6% (95%CI: 44.0-59.1), 63.6% (95%CI: 57.0-69.8), 62.5% (95%CI: 50.1-73.5), 63.6% (95%CI: 57.0-69.8), 62.5% (95%CI: 50.1-73.5), 74.7% (95%CI: 50.0-89.7), and 72.6% (95%CI: 58.1-83.5), respectively. The prevalence of elevated interleukin-6 was 59.9% (95%CI: 48.2-70.5), CD3 was 68.3% (95%CI: 50.1-82.2), reduced CD4 was 62.0% (95%CI: 51.1-71.6), reduced CD8 was 42.7% (95%CI: 32.2-53.9). The prevalence of elevated troponin-I was 20.6% (95%CI: 9.0-40.5), elevated creatine kinase-MB (CKMB) was 14.7% (95%CI: 7.1-28.0), elevated brain natriuretic peptide (BNP) was 48.9% (95%CI: 30.4-67.7), elevated blood urea nitrogen was 13.1% (95%CI: 6.6-24.4),, elevated creatinine was 7.2% (95%CI: 4.4-11.8), elevated lactate dehydrogenase (LDH) was 53.1% (95%CI: 43.6-62.4), hyperglycemia was 41.1% (95% CI: 28.2-55.5), elevated total bilirubin was 48.9% (95%CI: 30.4-67.7), reduced albumin was 54.7% (95%CI: 38.1-70.2), reduced pre-albumin was 49.0% (95%CI: 26.6-71.8), and reduced PT was 53.1% (95% CI: 43.6-62.4), and D-dimer was 44.9% (95%CI: 31.0-59.6).

**Conclusion:** This study provides a comprehensive description of laboratory characteristics in patients with COVID-19. The results show that lymphopenia, elevated CRP, elevated ESR, elevated ferritin, elevated serum amyloid A, elevated BNP, reduced albumin, reduced pre-albumin, reduced CD3, reduced CD4, reduced CD8, elevated D-dimer, reduced PT, elevated interleukin-2, elevated interleukin-6, elevated LDH and hyperglycemia are the common findings at the time of admission.

## 1. Background

Coronavirus disease 2019 (COVID-19) is a novel infectious disease with rapid human-to-human transmission and diverse mortality due to acute respiratory distress syndrome (ARDS), multiple organ failure, and other serious complications (1-3). In early December 2019, the first patient with COVID-19 pneumonia was found in Wuhan, Hubei Province, China (4). Since then, COVID-19 pneumonia has spread rapidly throughout China. Soon after, many countries announced the first case of COVID-19 pneumonia (5-7).

Severe acute respiratory syndrome coronavirus 2 (SARS-CoV-2) is the seventh identified coronavirus with human infection capacity. On the one hand, most human coronavirus infections are mild. On the other hand, coronavirus infections, including COVID-19, severe acute respiratory syndrome coronavirus (SARS-CoV) and also Middle East respiratory syndrome coronavirus (MERS-COV) can be severe or even fatal, especially in elderly patients and patients with hypertension, diabetes and renal failure (8, 9).

On January 30, 2020, the world health organization (WHO) announced the outbreak of the disease as public health emergency of international concern (PHEIC), because these uncertainties lead to the ongoing global transmission of the virus, and the need to track and control the outbreak in the world (10). According to a recent report, common symptoms include fever, fatigue, and dry cough (11).

Recent studies have suggested the role of primary laboratory tests in addition to clinical symptoms for suspected patients, which play a significant role in the diagnosis of COVID-19. But the results of these studies are contradictory (1, 2, 11-60). Given that researchers and practitioners face enormous amounts of information, systematic reviews reduce the levels of bias and random errors through a structured review of the literature. A meta-analysis is a statistical analysis that combines the results of multiple scientific studies. Meta-analysis can be performed when there are different scientific articles addressing the same question, with each individual study reporting measurements that are expected to have some level of error. The purpose is to use approaches from statistics to achieve a pooled estimate closest to the unknown truth based on how this error is perceived (61, 62). Therefore, the present study was conducted to evaluate laboratory findings in COVID-19 patients.

## 2. Method

### 2.1. Protocol

The present meta-analysis was reported in accordance with the Preferred Reporting Items for Systematic Reviews and Meta-analysis (PRISMA) guidelines (63). This protocol is registered with the code CRD42019145410 in PROSPERO International Database. All steps were performed by two authors (M.A and A.J.) and disagreements between the two authors regarding the quality of studies or extracted data were resolved after discussion with the senior author (M.K.).

### 2.2. Literature review

We did a comprehensive literature review in databases PubMed/Medline, Scopus, EMBASE, Web of Science, Cochrane Library (Cochrane Database of Systematic Reviews - CDSR), Ovid, Science Direct, CINAHL and EBSCO to find citations from the beginning of January 2019 to the beginning of April 2020 without any restrictions. A search for gray literature was conducted at https://www.medrxiv.org/. To ensure a thorough systematic literature review, we reviewed the reference lists of eligible studies and systematic eligible surveys identified from the above sources.

The search terms included “Novel coronavirus”, “COVID-19”, “2019 nCoV”, “Novel coronavirus 2019”, “acute respiratory infection”, “Wuhan pneumonia”, “Wuhan coronavirus”, “SARS-CoV-2”, “Laboratory”, “Laboratories”, “clinical features”, and “Clinical Characteristics”.

The following is a sample of the search in PubMed: (Novel coronavirus OR COVID-19 OR 2019 nCoV OR Novel coronavirus 2019 OR acute respiratory infection OR COVID-19 OR Wuhan pneumonia OR Wuhan coronavirus OR SARS-CoV-2) AND (Laboratory OR Laboratories OR clinical features OR Clinical Characteristics).

### 2.3. Inclusion and exclusion criteria

Studies with the following criteria were included: 1) All cross-sectional studies examining laboratory findings and 2) the studies with full test. Studies with the following criteria were excluded: 1) the sampling method was non-random, 2) duplicate studies, 3) lack of relevance to the subject, 4) diagnostic intervention for COVID-19 other than laboratory confirmation, 5) sample size of less than 10 participants, 6) low quality in the qualitative evaluation and 8) case studies, review articles, letter to the editor without quantitative data.

### 2.4. Data extraction

Extracted data include: author’s name and year of publication, country and province, study source, study design, total sample size, number of men and women, mean age and standard deviation, patient description, number of patients referred to the intensive care unit (ICU), COVID-19 diagnostic method, test sample for COVID-19 diagnosis (respiratory secretions, blood, etc.), sample location (nasal, pharyngeal), and available laboratory findings (including CBC components, inflammatory markers, cellular immunity tests, cytokines, and cardiac, renal, hepatic, muscular, and coagulopathic tests). In this study, only the number of cases that increased or decreased in each test was extracted based on normal range, and the mean and standard deviation of the experiments were not extracted.

If two articles were published by the same authors or by the same institute, one of those studies was selected with a bigger sample size or the information of both articles was used.

### 2.5. Qualitative evaluation

Critical evaluation of studies was performed using the Newcastle-Ottawa Scale (NOS) (64). Studies with six stars or more (based on a scale of 0 to 9) were considered moderate to good quality studies. In terms of study quality, studies with scores 6-7 were considered as average quality and studies with scores 8-9 were considered as good quality.

### 2.6. Statistical analysis

In the present meta-analysis, the total number and positive cases for each test were used. We tested the statistical heterogeneity between the studies using I^2^ and Q statistic; I^2^>50% and P<0.10 were statistically significant (65). In the case of significant heterogeneity, we proposed meta-analysis based on random effects model, because this model can estimate the distribution of true effect size (ES) (66). Sensitivity analysis was also performed by omitting one study at a time and combining the results of the remaining studies. We did the meta-analysis in Comprehensive Meta-Analysis Software (CMA) version 2. The publication bias was assessed using the Begg’s test and Egger’s test for most variables with the highest number of studies (67, 68).

## 3. Results

### 3.1. Search results and study features

Figure 1 shows the flowchart for the selection of studies. In the electronic search, 2652 related topics were retrieved. Articles were reviewed based on their title and abstract, and 821 duplicate and 1682 unrelated articles were deleted. Ninety-seven studies were removed after a full review of the text because they did not meet the criteria for entering the study (Figure 1). Finally, 52 studies entered the meta-analysis process after qualitative evaluation. All studies were conducted in China and all of them were of good quality (Table 1). The average age of the participants was estimated to be 52.09 years (95% CI: 48.80-55.38).

**Figure 1:**
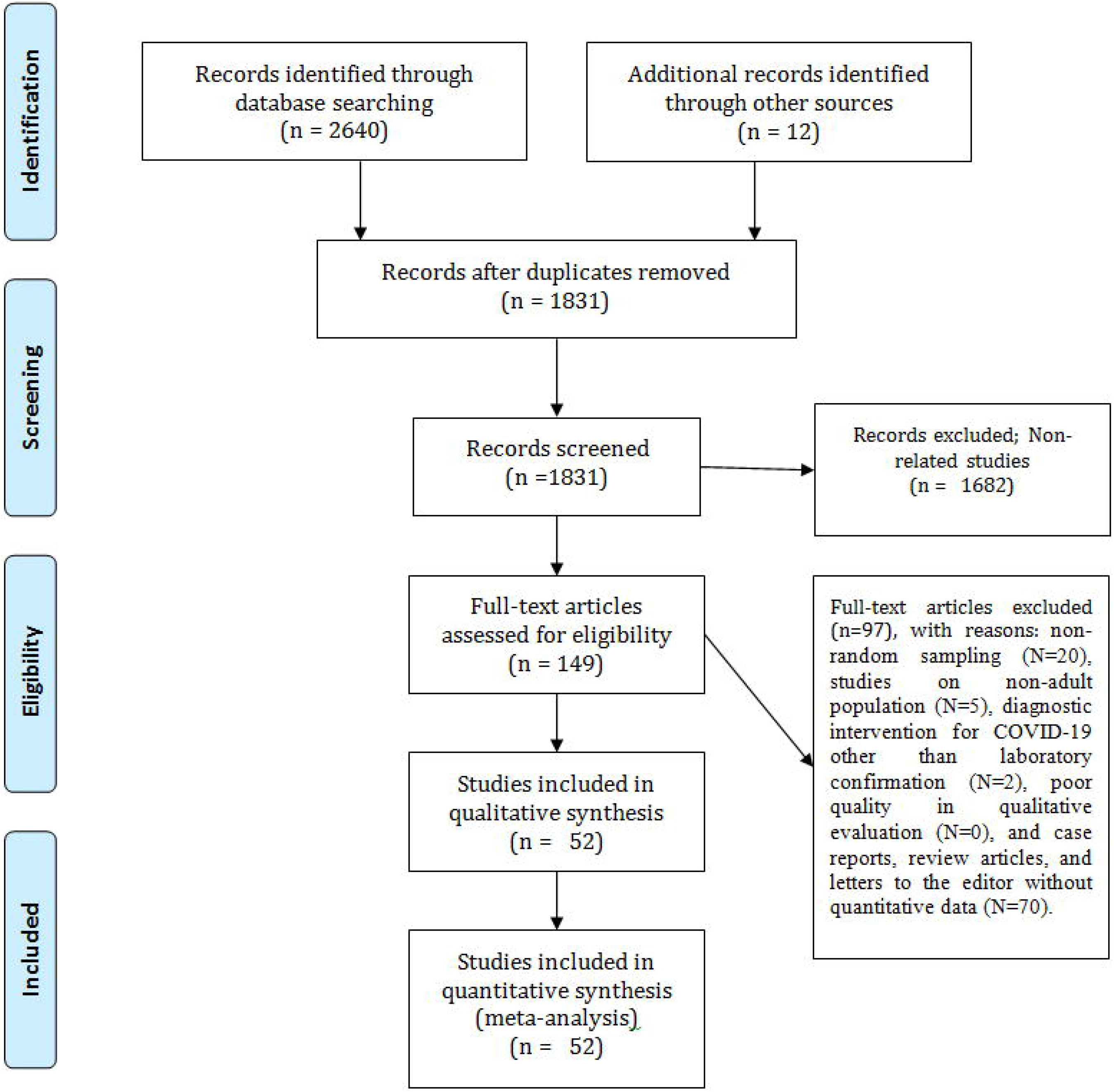
PRISMA flowchart

### 3.2. Complete blood count components

The prevalence of leukocytosis, lymphocytosis, neutrophilia, and monocytosis in patients with COVID-19 was estimated to be 9.9% (95% CI: 7.7-12.7), 4.0% (95% CI: 1.3-11.4), 18.9% (95% CI: 13.1-26.6), and 17.1% (95% CI: 127-22.6), respectively (Figure 2A-D).

**Figure 2:**
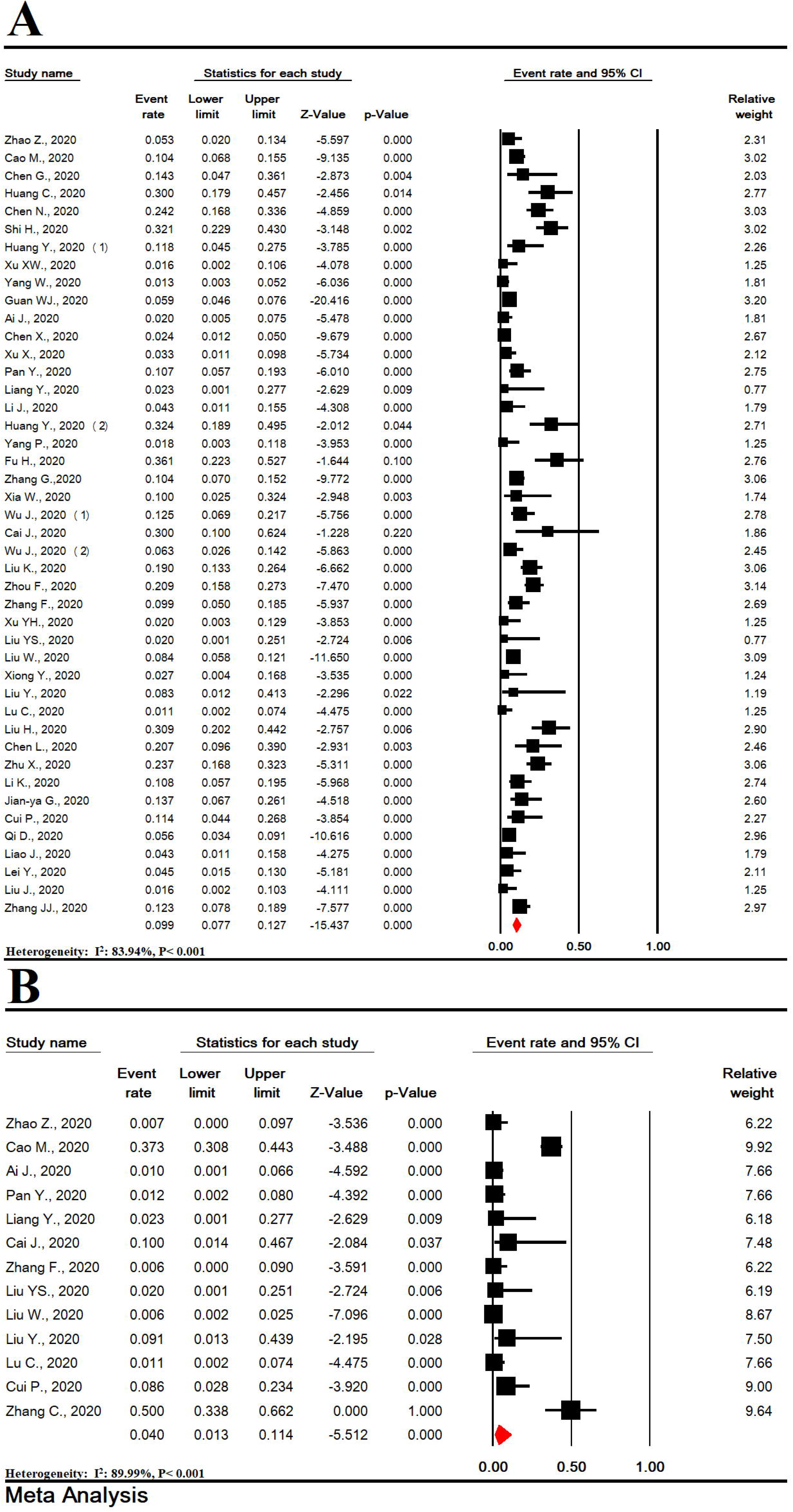
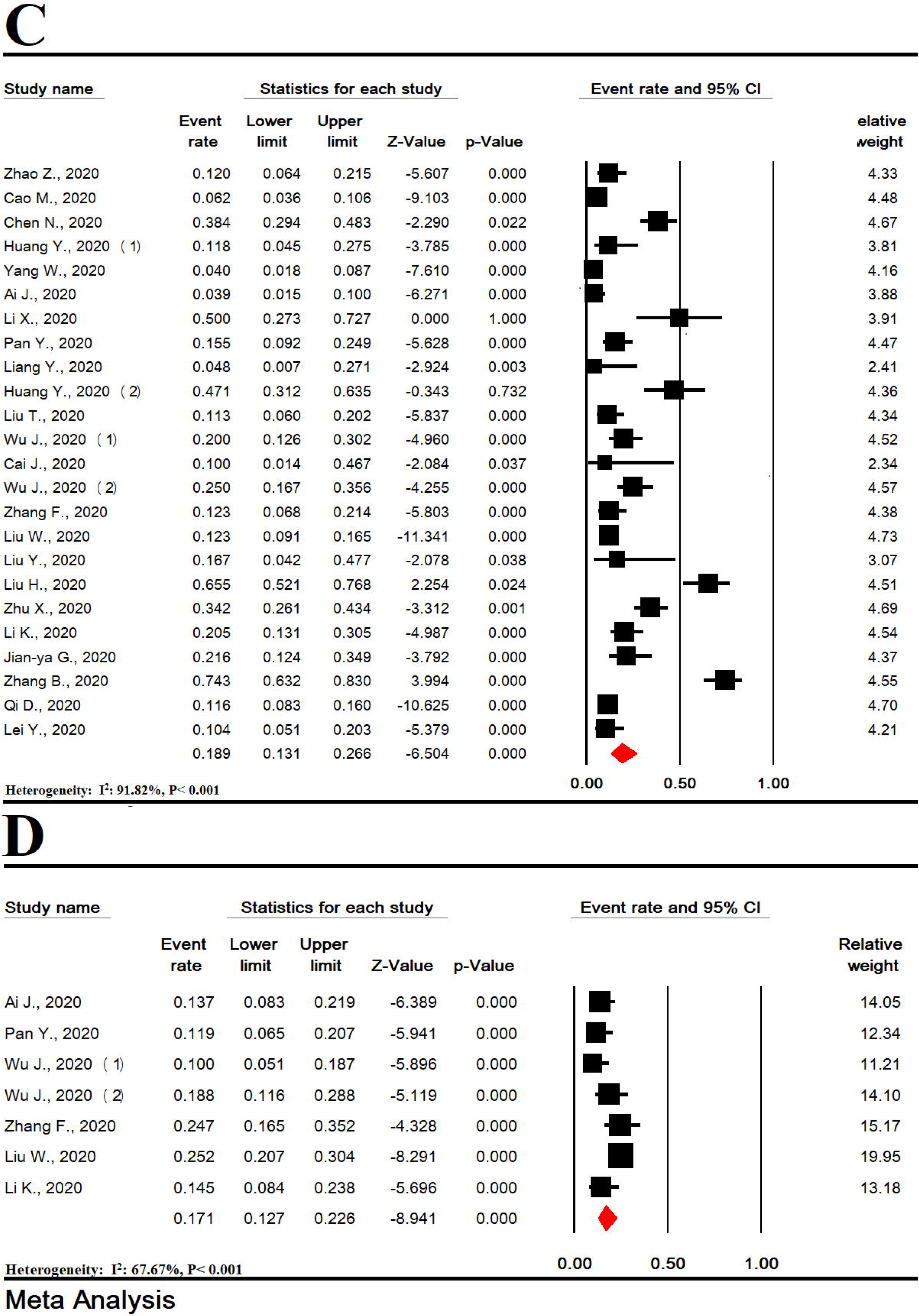
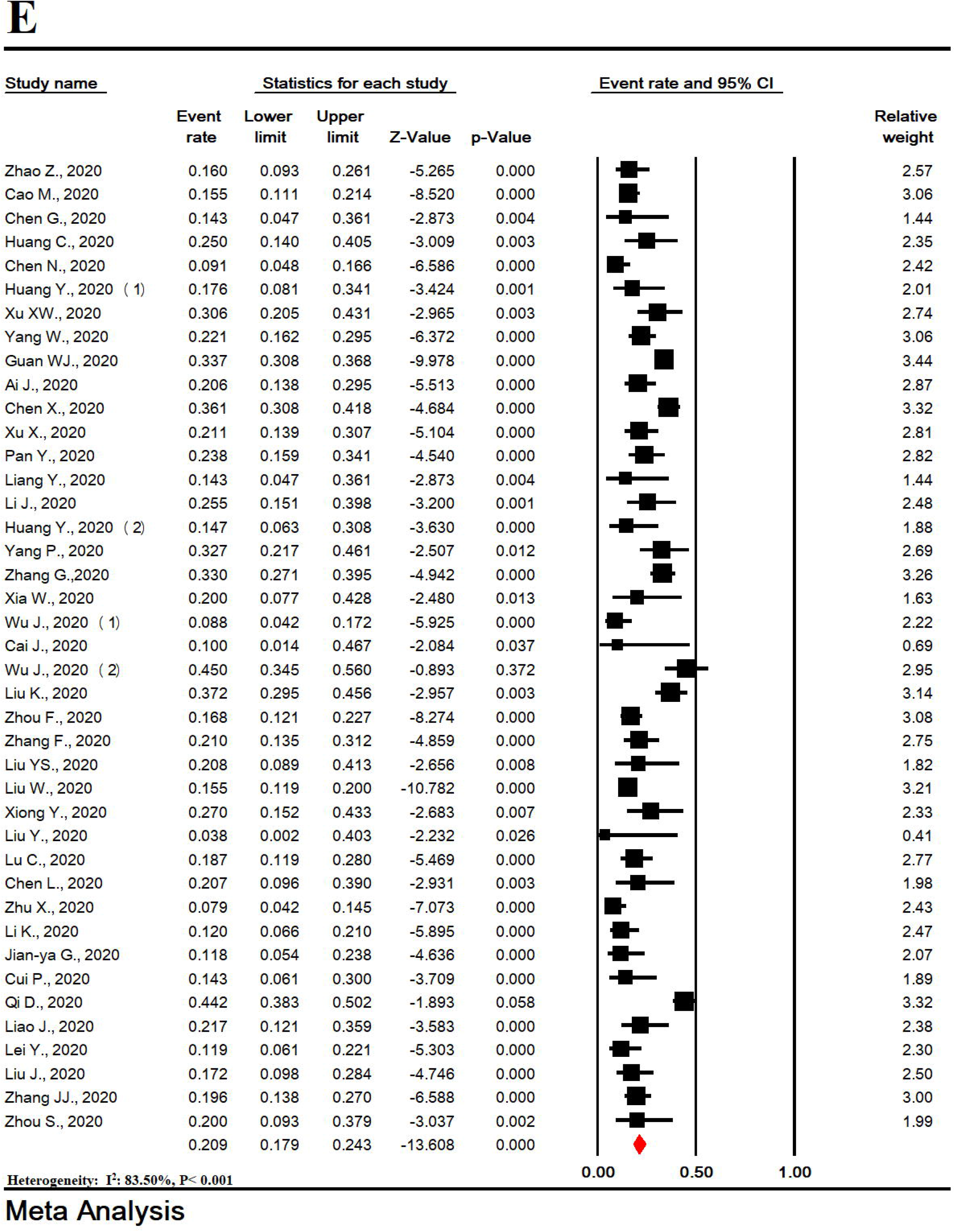
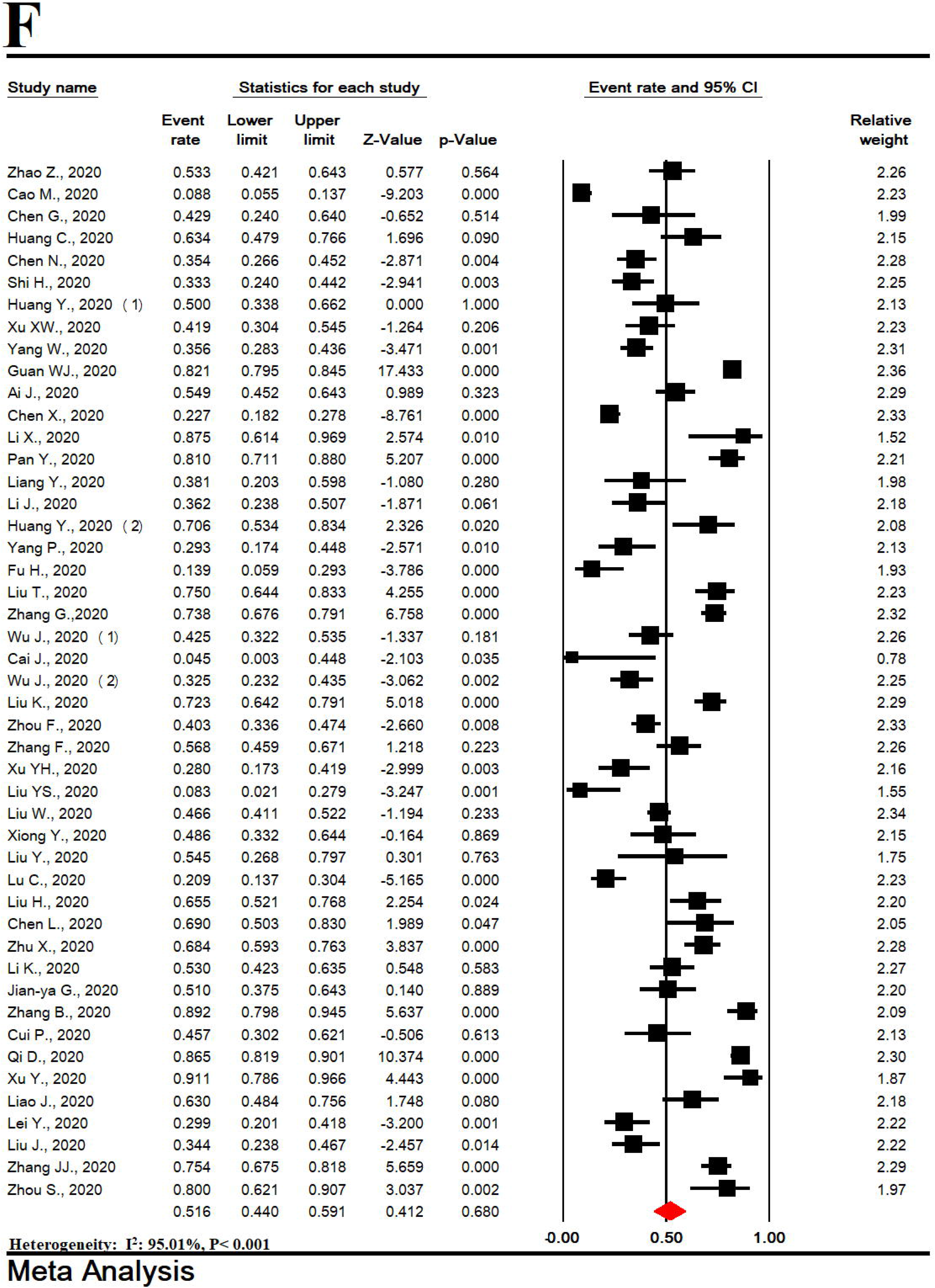
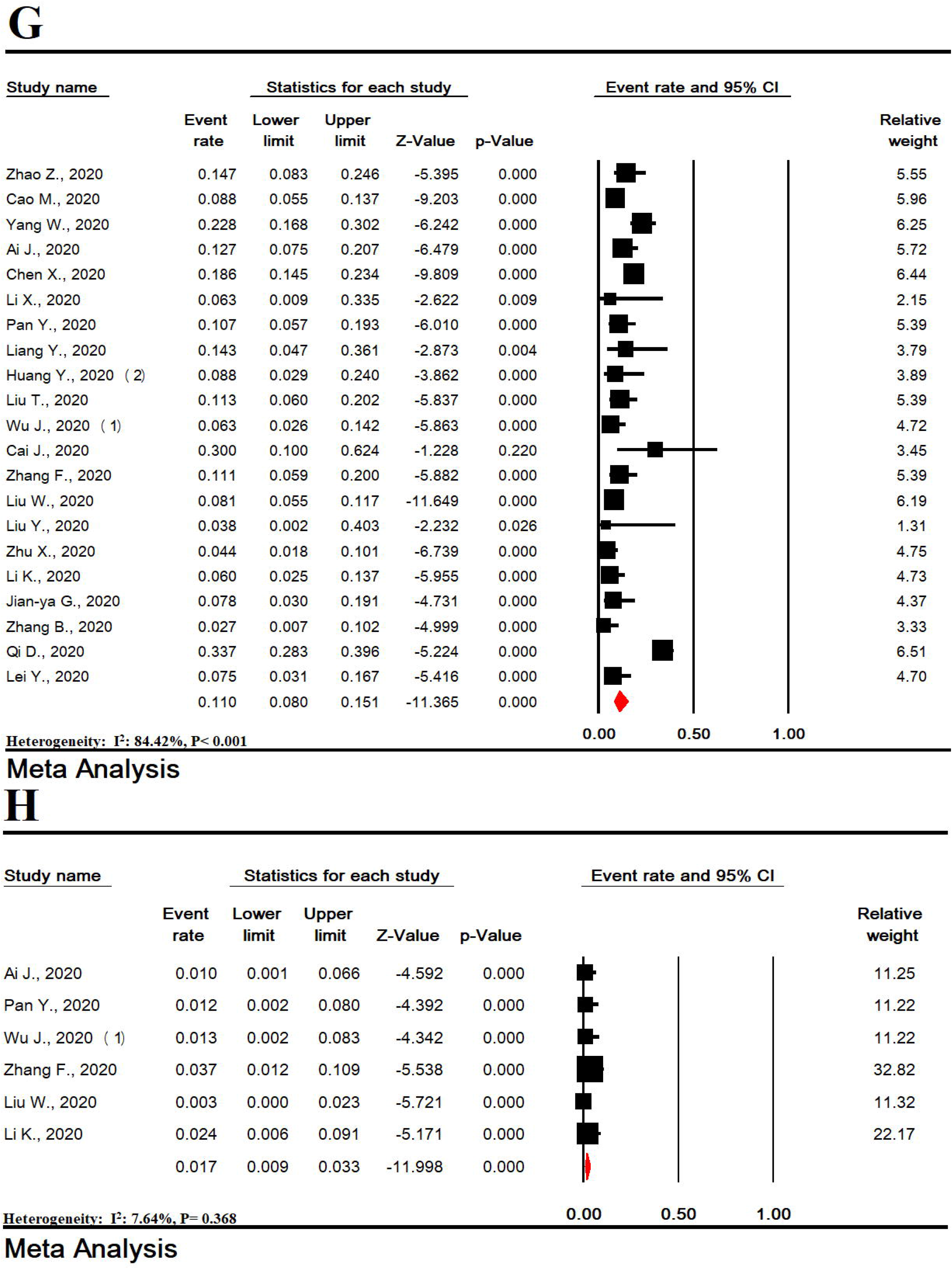
The prevalence of leukocytosis (A), lymphocytosis (B), neutrophilia (C), monocytosis (D), leucopenia (E), lymphopenia (F), neutropenia (G) and monocytopenia (H) in patients with COVID-19.

The prevalence of leukopenia, lymphopenia, neutropenia and monocytopenia in patients with COVID-19 was estimated to be 20.9% (95% CI: 17.9-24.3), 51.6% (95% CI: 44.0-59.1), 11.0% (95% CI: 8.0-15.1), 1.7% (95% CI: 0.9-3.3), respectively (Figure 2E-I).

The prevalence of thrombocytosis was 4.7% (95% CI: 2.3-9.1) and thrombocytopenia was 17.2% (95% CI: 13.7-21.5) (SF 2A-B). The prevalence of polycythemia was 26.5% (95% CI: 15.7-41.2) and anemia 3.3% (95% CI: 0.3-27.4) (SF 2 C-D).

### 3.3. Inflammatory markers

The prevalence of elevated c-reactive protein (CRP), elevated erythrocyte sedimentation rate (ESR), elevated TNFα, elevated serum amyloid A, elevated ferritin and elevated procalcitonin (PCT), in patients with COVID-19 was estimated to be 63.6% (95% CI: 57.0-69.8), 62.5% (95% CI: 73.5-50.1), 28.7% (95% CI: 9.0-62.1), 74.7% (95% CI: 50.0-89.7), 19.1% (95% CI: 13.1-26.9), and 72.6% (95% CI: 58.1-83.5),. respectively (Figure 3A-F).

**Figure 3:**
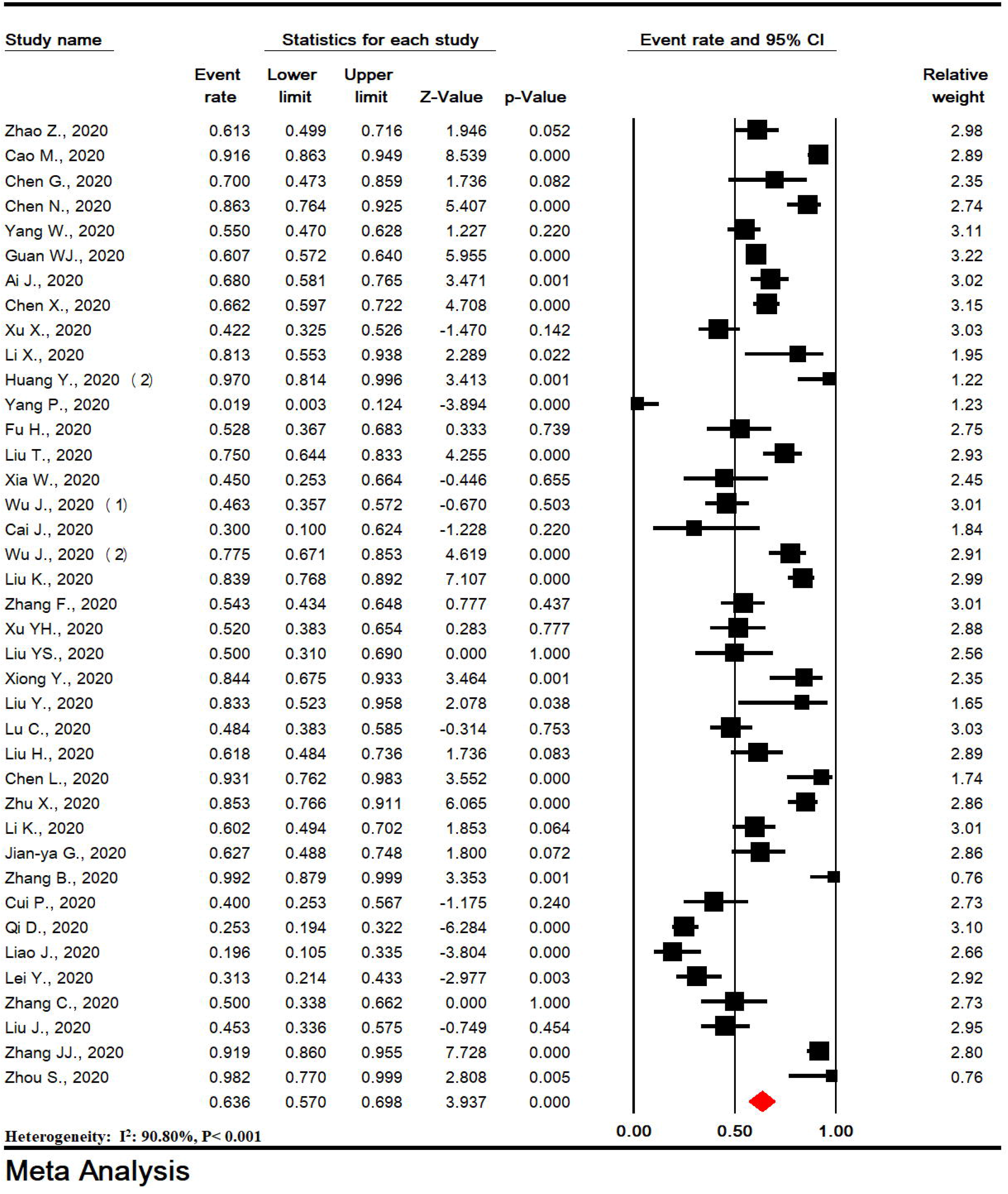
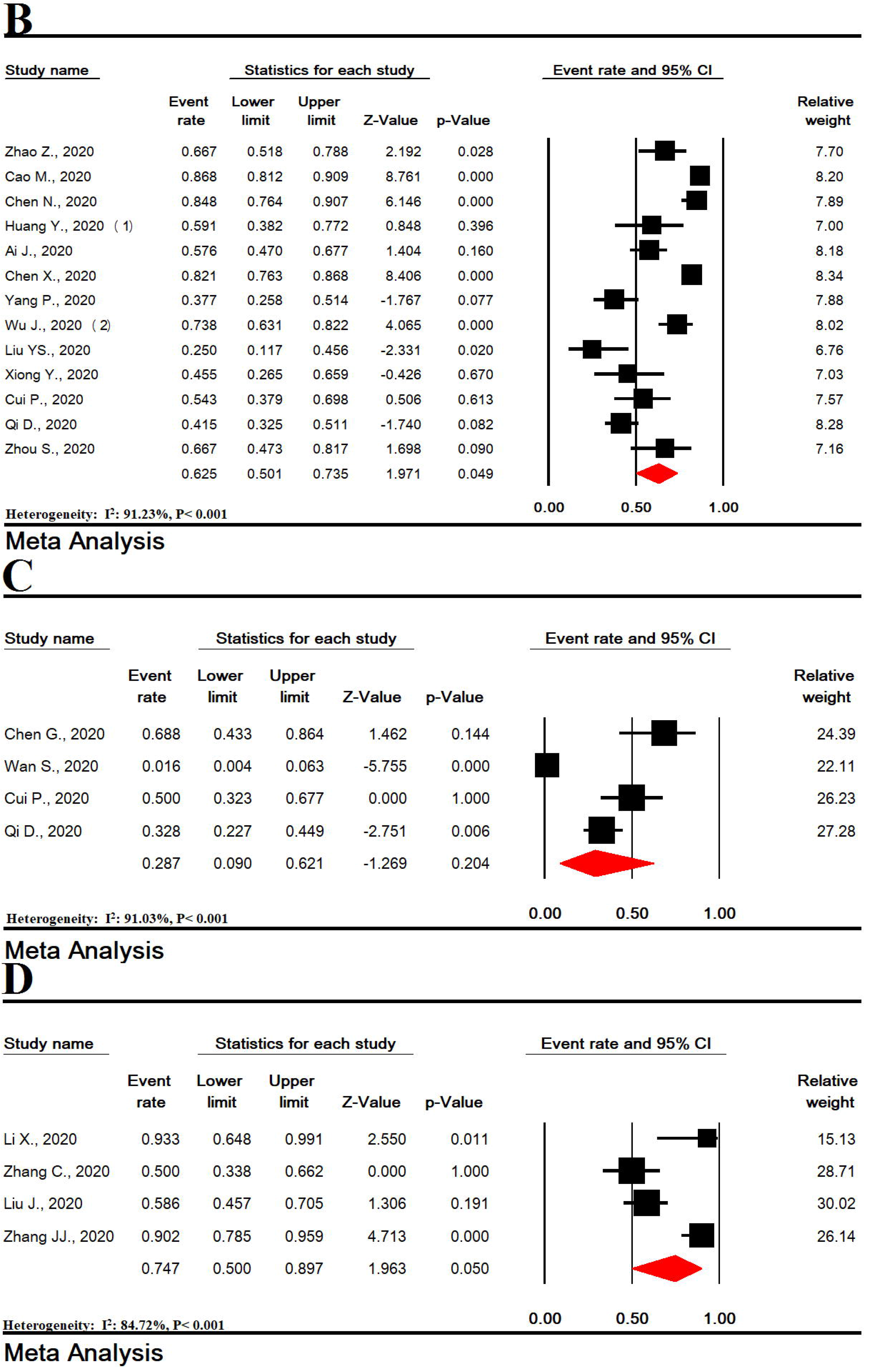
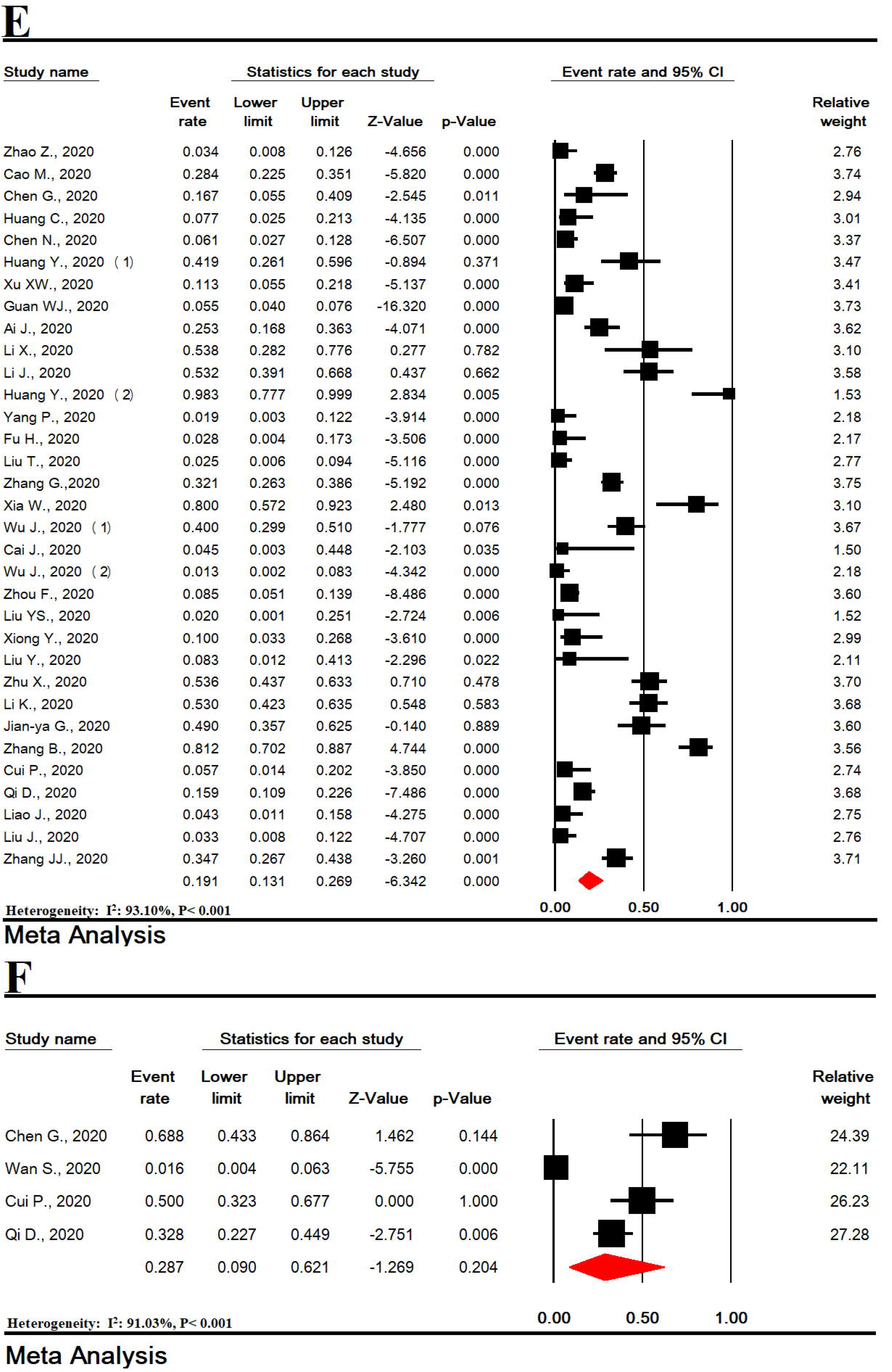
The prevalence of elevated C - reactive protein (A), elevated erythrocyte sedimentation rate (B), elevated TNFα (C), elevated serum amyloid A (D), elevated procalcitonin (E), and elevated ferritin (F) in patients with COVID-19.

### 3.4. Cytokines

In patients with COVID-19, the prevalence of elevated interleukin-2 was 32.0% (95% CI: 8.0-71.8), elevated interleukin-6 was 59.9% (95% CI: 48.2-70.5) and elevated interleukin-10 was 12.7% (95% CI: 2.9-41.8) (Figure 3A-C).

### 3.5. Cell immunity testes

The prevalence of reduced CD3, reduced CD4, reduced CD8, reduced CD4/CD8 and elevated CD4/CD8 in patients with COVID-19 was estimated to be 68.3% (95% CI: 50.1-82.2), 62.0% (95% CI: 51.1-71.6) and 42.7% (95% CI: 32.2-53.9), 9.3% (95% CI: 5.6-15.2), and 15.3% (95% CI: 4.6-39.9), respectively (SF3 A-E).

### 3.6. Cardiac tests

In patients with COVID-19, the prevalence of elevated troponin-I (cTnI) was 20.6% (95% CI: 9.0-40.5), elevated creatine kinase-MB (CKMB) was 14.7% (95% CI: 7.1-28.0), elevated brain natriuretic peptide (BNP) was 48.9% (95% CI: 30.4-67.7) (SF 4).

### 3.7. Kidney tests

The prevalence of elevated blood urea nitrogen (BUN), elevated creatinine, hyperkalemia and hypokalemia in patients with COVID-19 was estimated to be 13.1% (95% CI: 6.6-24.4), 7.2% (95% CI: 4.4-11.8), 15.1% (95% CI: 8.1-26.4), and 15.6% (95% CI: 12.8-19.0), respectively (SF 5).

### 3.8. Liver tests

In patients with COVID-19, the prevalence of elevated alanine aminotransferase (ALT) was 20.6% (95% CI: 9.0-40.5), elevated aspartate aminotransferase (AST) was 14.7% (95% CI: 7.1-28.0), elevated lactate dehydrogenase (LDH) was 53.1% (95% CI: 43.6-62.4), hyperglycemia was 41.1% (95% CI: 28.2-55.5), elevated total bilirubin was 48.9% (95% CI: 30.4-67.7), elevated direct bilirubin was 14.7% (95% CI: 7.1-28.0) and elevated globulin was 5.3% (95% CI: 1.8-14.5) (SF 6-7). The prevalence of reduced albumin and reduced pre-albumin in patients with COVID-19 was estimated to be 54.7% (95% CI: 38.1-70.2) and 49.0% (95% CI: 26.6-71.8), respectively (SF 7).

### 3.9. Muscle tests

In patients with COVID-19, the prevalence of elevated of myoglobin was 18.3% (95% CI: 9.9-31.5) and elevated creatine phosphokinase (CPK) was 13.7% (95% CI: 11.0-16.9) (SF 8).

### 3.10. Coagulation tests

The prevalence of elevated D-dimer, elevated prothrombin time (PT), elevated partial thromboplastin time (PTT) in patients with COVID-19 was estimated to be 44.9% (95% CI: 31.0-59.6), 15.2% (95% CI: 5.9-33.7) and 20.6% (95% CI: 11.3-34.6), respectively (Figure 5) (SF 9 A-B).

**Figure 4:**
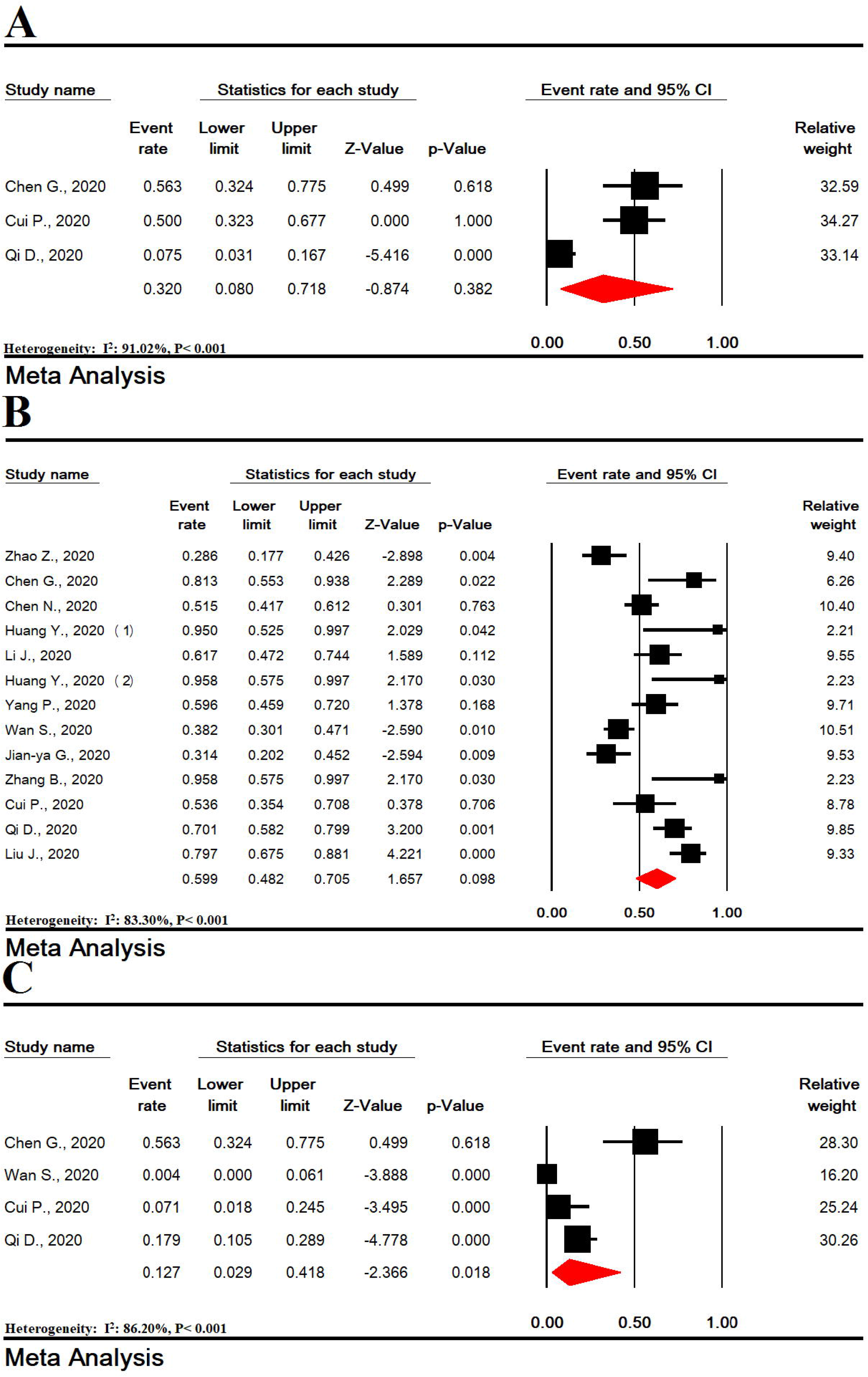
The prevalence of elevated interleukin-2 (A), elevated interleukin-6 (B) and elevated interleukin-10 (C) in patients with COVID-19.

**Figure 5:**
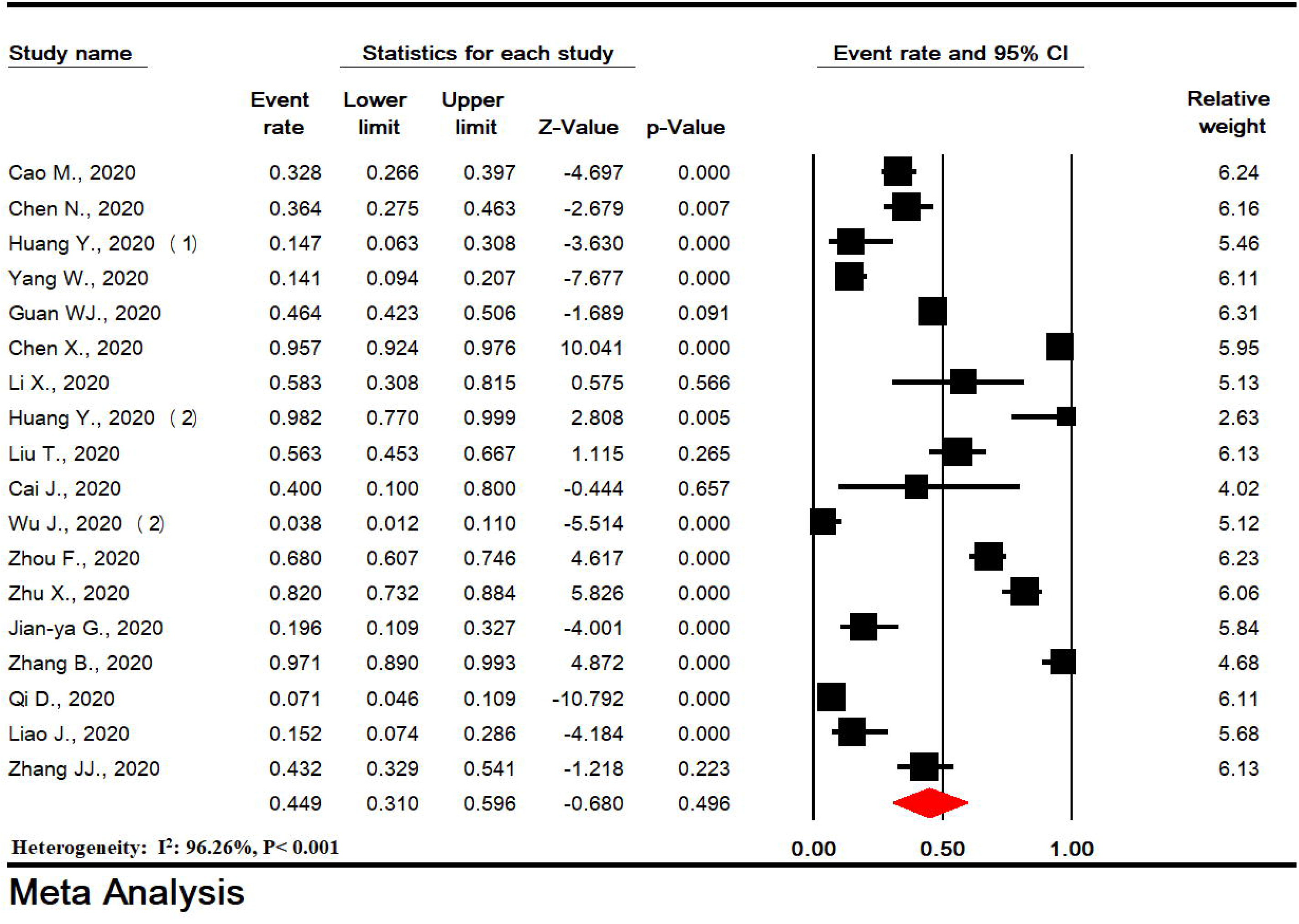
The prevalence of D-dimer in patients with COVID-19.

In patients with COVID-19, the prevalence of reduced PT was 53.1% (95% CI: 43.6-62.4) and reduced PTT was 4.1% (95% CI: 1.1-14.5) (SF 9 C-D).

### 3.11. Sensitivity analysis

Sensitivity analysis for all meta-analyses showed that the overall result is robust even after the elimination of one individual study (SF 10 A-GG).

### 3.12. Publication bias

Publication bias for performed meta-analyses showed that publication bias had no effect on studies. Begg’s test and Egger’s test results were as follows: Egger = 0.115 and Begg = 0.840 for lymphocytosis, Egger = 0.307 and Begg = 0.199 for CRP increase, Egger = 0.155 and Begg = 0.220 for procalcitonin increase, and Egger = 0.779 and Begg = 0.186 for AST increase (SF 11).

## 4. Discussion

Our systematic review and meta-analysis included 52 studies and 5490 patients, which provides the most comprehensive overview of the laboratory findings about patients with COVID-19. Compared to previous systematic studies published on this subject, more studies and laboratory results are covered in the present analysis (69, 70).

In the present meta-analysis, the most common estimated laboratory abnormalities included Lymphopenia (51.6%), elevated CRP (63.6%), elevated ESR (62.5%), elevated ferritin (72.6%), elevated serum amyloid A (74.7%), elevated BNP (48.9%), reduced albumin (54.7%), reduced pre-albumin (49.0%), reduced CD3 (68.3%), reduced CD4 (62.0%), reduced CD8 (42.7%), elevated D-dimer (44.9%), reduced PT (53.1%), elevated interleukin-2 (32.0%), elevated Interleukin-6 (59.9%), elevated LDH (53.1%) and hyperglycemia (41.1%).

However, all of these laboratory markers are non-specific, which limits their clinical use. When assessing suspected cases, physicians cannot rely on these laboratory abnormalities to reject or confirm the diagnosis of COVID-19, and they only play a supporting role. These abnormalities are similar to those previously seen in patients with SARS and MERS (71-73).

In this meta-analysis, changes in lymphocytes predominantly included a decrease in lymphocytes. Pathological results in patients with COVID-19 showed that overactive T lymphocytes, characterized by increased Th17 cells and high toxicity of CD8 + T cells, caused some severe immune damage. This may be the main reason for the disappearance of lymphocytes in these patients (74, 75).

In the present meta-analysis, a decrease in CD3, CD4, and CD8 cells was observed in most COVID-19 patients. CD4 + T cells play an important role in regulating immune responses, cell removal and strengthening of immune cells, especially CD8 + T cells (76). CD4 + T cells facilitate the production of specific antibodies to eradicate the virus by activating T-dependent B cells. On the other hand, CD8 + T cells exert their effects mainly through two mechanisms, including cytolytic activity against target cells or secretion of cytokines, including IFg, TNFα, and IL-2, as well as many chemokines (77). However, continuous stimulation by the virus may cause T cell exhaustion, leading to loss of cytokine production capacity and decreased function (78, 79). Other studies have shown that the number of T cells in patients with COVID-19 is significantly reduced, and the surviving T cells appear to be worn out. Studies also showed that in non-ICU patients, complete T cells, CD8 + T cells and CD4 + T cells were less than 800 microliters, 300 microliters and 400 microliters, respectively. More invasive interventions may be required even if severe symptoms are not observed. That’s because they are at high risk for worse conditions (80).

Since there is no proven antiviral treatment yet, there may be ways to boost the immune system. On the other hand, the most common hematological changes in COVID-19 patients are lymphopenia and immune system defects. We assume that hematopoietic growth factors such as G-CSF, which mobilize endogenous blood stem cells and endogenous cytokines, may be a possible hematological treatment for COVID-19 patients (81).

In the present study, inflammatory markers such as CRP, ESR, ferritin, PCT, and serum amyloid A were increased in most patients. CRP is a non-specific acute-phase reactive protein derived from IL-6 in the liver and is a biomarker sensitive to inflammation, infection, and tissue damage (82). CRP expression levels are usually low, but increase sharply and significantly during acute inflammatory responses (68, 83). Increased CRP in isolation or in combination with other markers may indicate bacterial or viral infections. Other studies have shown that in the early stages of COVID-19, CRP levels are positively associated with lung lesions and can reflect the severity of the disease (84, 85). PCT is a glycoprotein without hormonal activity and is an important calcitonin (85, 86). Serum PCT levels are usually low or undetectable(87). PCT levels are increased by bacterial infections and are relatively low with viral infections. Therefore, they can be used to differentiate between bacterial and viral infections (88). Higher PCT levels in the severe COVID-19 group indicate that critically ill patients may have concomitant bacterial infections (89).

In a recent meta-analysis on patients with COVID-19-induced ARDS, physicians were advised to closely monitor the number of WBCs, the number of lymphocytes, the number of platelets, IL-6, and serum ferritin as a sign of potential progression to critical illness. PCT should also be measured regularly to act as an indicator of secondary bacterial infection, which is frequently seen in patients who do not survive. Finally, these findings should be continuously reviewed in the upcoming months (59).

In the present meta-analysis, abnormalities in liver tests such as AST, ALT, LDH, bilirubin, globulin, albumin and pre-albumin were observed in COVID-19 patients. The liver has bile receptors, and these abnormalities can be the result of bile damage (33, 90). This could be an explanation for the liver’s abnormal laboratory findings in the early stages of infection. Hypoxia is a serious event in COVID-19 and is one of the leading causes of sudden death in patients, so an increase in CPK may be the result of hypoxia and should be causally interpreted (59).

The similarity between DNA sequencing of the COVID-19 and SARS-COV suggests that the mechanism of action may be similar (91). SARS–COV enters the host cells through the S-spike protein, acts on the bronchial epithelial cells through the ACE2 receptor, and then infects other cells. In severe cases, it can cause a series of immune responses or cytokine storms (92, 93). In our meta-analysis, cytokines, including Interleukin-2 and Interleukin-6, were elevated in most patients. Cytokine storms are a phenomenon of acute inflammatory reaction in which cytokines are rapidly produced in response to a large number of microbial infections. This phenomenon plays an important role in ARDS and multiple organ dysfunction syndrome (MODS) (94, 95). It has also been involved in the regulation of respiratory viral infections such as SARS viral infection in 2002, and H7N9 viral infection in 2013 (96).

In one meta-analysis, cTnI analysis in four studies showed that the levels of cTnI in patients with severe SARS-CoV-2 infection increased compared to those with milder forms of the disease. It is therefore reasonable to assume that the initial measurement of cardiac injury markers in SARS-CoV-2 patients immediately after hospitalization, as well as long-term monitoring during hospitalization, may help identify patients with possible cardiac injury. This way, it is possible to predict the progression of COVID-19 towards worse clinical conditions (97).

In the present study, elevated levels of creatinine, hyperkalemia and hypokalemia were observed in patients with COVID-19. The exact mechanism of kidney involvement is unknown. The hypothetical mechanism includes sepsis, which leads to cytokine storm syndrome or direct cell damage caused by the virus. Angiotensin converting enzyme (ACE) and dipeptidyl peptidase IV (DPP-IV) have both been expressed in renal tubular epithelial cells, and have been identified as a connecting partner for SARS-CoV and MERS-CoV, respectively (98). Viral RNA has been identified in kidney tissue and urine in both infections (72). Recently, Zhong’s lab in Guangzhou successfully isolated SARS-CoV-2 from the urine sample of an infected patient and identified the kidney as the target of this novel virus. Previous reports of SARS and MERS-CoV infections have shown acute kidney injury (AKI) in 5% to 15% of cases and high mortality rates (60-90%). Preliminary reports indicate a lower prevalence (9%) of AKI in people with COVID-19 infection (1, 17, 55). AKI was an independent risk factor for hospital mortality (99).

Recent literature shows that D-dimer values are often increased in patients with COVID-19, which was reported to be 44.9% in the present meta-analysis. What is clear is that D-dimer values are severe in patients with COVID-19, even in patients with milder forms, and therefore, D-dimer measurement may be associated with a worsening clinical picture. Although an increase in D-dimer is known to be a multifactorial cause, Lippi et al. stated that an increase in D-dimer and disseminated coagulopathy might be a trivial finding in patients with severe COVID-19 forms compared to other infectious diseases such as human immunodeficiency virus (HIV), Ebola, Zica virus, and Chikungunya virus. Therefore, immediate studies should be planned to determine whether antithrombotic adjunctive treatments (e.g., anticoagulants, antithrombin, or thrombomodulin) may be helpful in patients with severe COVID-19 (100).

In fact, it has been reported that coagulation is activated and accelerated in response to several infections because this mechanism may enhance the physiological response (101-103). Coagulation also has immune function, which can be another line of defense against severe infection (104). Although it is reasonable to assume that homeostasis is deviated in patients with COVID-19, overuse of coagulation factors increases the risk of disseminated intravascular coagulation (DIC), which has a clear negative effect on prognosis (105). A study by Han et al. found that coagulation in COVID-19 patients was clearly deranged compared to a healthy control population (106).

The limitations of this study include the following: 1) All studies were conducted in China and laboratory manifestations may be influenced by ethnic factors. 2) Most patients are hospitalized and patients who have milder symptoms or patients who are not hospitalized may cause bias in the results. 3) In most early studies, laboratory findings were not considered separately for patients admitted to the intensive care unit or the isolated ward. Our results are based on laboratory findings at the time of admission, but patients may have experienced symptoms before admission (because laboratory findings are influenced by the clinical course of the disease), and during this time, the patient may receive antiviral or antibacterial drugs. 4) Since all studies were performed in China in 2020 using the similar diagnostic method, we were unable to determine the cause of the heterogeneity.

## Conclusion

This study provides a comprehensive description of laboratory characteristics in patients with COVID-19. The results show that lymphopenia, elevated CRP, elevated ESR, elevated ferritin, elevated serum amyloid A, elevated BNP, reduced albumin, reduced pre-albumin, reduced CD3, reduced CD4, reduced CD8, elevated D-dimer, reduced PT, elevated interleukin-2, elevated interleukin-6, elevated LDH and hyperglycemia are the common findings at the time of admission.

## Data Availability

Not applicable.

## Abbreviations

COVID-19: Coronavirus disease 2019
ARDS: Acute respiratory distress syndrome
SARS-CoV-2: Severe acute respiratory syndrome coronavirus 2
SARS-CoV: Severe acute respiratory syndrome coronavirus
MERS-COV: Middle East respiratory syndrome coronavirus
WHO: world health organization
PHEIC: Public health emergency of international concern
PRISMA: Preferred Reporting Items for Systematic Reviews and Meta-analysis
CI: Confidence interval
ICU: intensive care unit
NOS: Newcastle-Ottawa Scale
CMA: Comprehensive Meta-Analysis Software
CRP: c-reactive protein
ESR: erythrocyte sedimentation rate
PCT: procalcitonin
cTnI: troponin-I
CKMB: creatine kinase-MB
BNP: brain natriuretic peptide
BUN: blood urea nitrogen
ALT: alanine aminotransferase
AST: aspartate aminotransferase
LDH: lactate dehydrogenase
CPK: creatine phosphokinase
PT: prothrombin time
PTT: partial thromboplastin time
MODS: multiple organ dysfunction syndrome
ACE: Angiotensin converting enzyme
DPP-IV: dipeptidyl peptidase IV
AKI: acute kidney injury
DIC: Disseminated intravascular coagulation

## Declarations

### Ethics approval and consent to participate

Not applicable.

### Consent for publication

Not applicable.

### Availability of data and materials

Not applicable.

### Competing interests

We declare no competing interests.

### Funding

This study was funded by the Ilam University of Medical sciences. Funder role was only financial support.

### Authors’ contributions

MA, ARJ, GhB, and MK acquired the data. MA analyzed and interpreted the data. MA, ARJ, GhB, and MK drafted the manuscript; MA, ARJ, GhB, and MK critically revised the manuscript for important intellectual content. MK supervised the study. All authors have read and approved the manuscript.

## Acknowledgement

We sincerely thank Ilam University of Medical Sciences for helping us with this research.

## Finger legends

SF 1: PRISMA checklist.

SF 2: The prevalence of thrombocytosis (A), thrombocytopenia (B), polycythemia (C), and anemia (D) in patients with COVID-19.

SF 3: The prevalence of reduced CD3 (A), reduced CD4 (B), reduced CD8 (C), reduced CD4/CD8 (D) and elevated CD4/CD8 (E) in patients with COVID-19.

SF 4: The prevalence of elevated troponin-I (A), elevated Creatine kinase-MB (B), elevated brain natriuretic peptide (C) in patients with COVID-19.

SF 5: The prevalence of elevated *blood urea nitrogen (A)*, elevated creatinine (B), hyperkalemia (C) and hypokalemia (D) in patients with COVID-19.

SF 6: The prevalence of elevated alanine aminotransferase (A), elevated aspartate aminotransferase (B), elevated lactate dehydrogenase (C), hyperglycemia (D), elevated total bilirubin (E), elevated direct bilirubin (F) and elevated globulin (G) in patients with COVID-19.

SF 7: The prevalence of reduced albumin (A) and reduced pre-albumin (B) in patients with COVID-19.

SF 8: The prevalence of elevated of myoglobin (A) and elevated creatine phosphokinase (B) in patients with COVID-19.

SF 9: The prevalence of elevated prothrombin time (PT) (A), elevated partial thromboplastin time (PTT) (B), reduced PT (C) and reduced PTT (D) in patients with COVID-19.

SF 10: Sensitivity analysis the prevalence of for leukocytosis (A), lymphocytosis (B), neutrophilia (C), monocytosis (D), leucopenia (E), lymphopenia (F), neutropenia (G), C - reactive protein (H), elevated erythrocyte sedimentation rate (I), elevated procalcitonin (J), elevated interleukin-6 (K), D-dimer (L), thrombocytosis (M), thrombocytopenia (N), polycythemia (O), reduced CD3 (P), reduced CD4 (Q), reduced CD8 (R), reduced CD4/CD8 (S) and elevated CD4/CD8 (T), elevated *blood urea nitrogen (U)*, elevated creatinine (V), elevated alanine aminotransferase (W), elevated aspartate aminotransferase (X), elevated lactate dehydrogenase (Y), hyperglycemia (Z), reduced albumin (AA), myoglobin (BB) and elevated creatine phosphokinase (CC), elevated prothrombin time (DD), reduced PT (EE), elevated troponin-I (FF), and elevated Creatine kinase-MB (GG) in patients with COVID-19.

## Notes

### Competing Interest Statement

The authors have declared no competing interest.

## References

1 Chen N, Zhou M, Dong X, Qu J, Gong F, Han Y, et al. Epidemiological and clinical characteristics of 99 cases of 2019 novel coronavirus pneumonia in Wuhan, China: a descriptive study. The Lancet. 2020;395(10223):507–13.

2 Huang C, Wang Y, Li X, Ren L, Zhao J, Hu Y, et al. Clinical features of patients infected with 2019 novel coronavirus in Wuhan, China. The Lancet. 2020;395(10223):497–506.

3 Li Q, Guan X, Wu P, Wang X, Zhou L, Tong Y, et al. Early transmission dynamics in Wuhan, China, of novel coronavirus–infected pneumonia. New England Journal of Medicine. 2020.

4 Lu R, Zhao X, Li J, Niu P, Yang B, Wu H, et al. Genomic characterisation and epidemiology of 2019 novel coronavirus: implications for virus origins and receptor binding. The Lancet. 2020.74–565:(10224)395;

5 Phan LT, Nguyen TV, Luong QC, Nguyen TV, Nguyen Ht, L. Hq, et al. Importation and human-to-human transmission of a novel coronavirus in Vietnam. New England Journal of Medicine. 2020;382(9):872–4.

6 Rothe C, Schunk M, Sothmann P, Bretzel G, Froeschl G, Wallrauch C, et al. Transmission of 2019-nCoV infection from an asymptomatic contact in Germany. New England Journal of Medicine. 2020;382(10):970–1.

7 Holshue ML, DeBolt C, Lindquist S, Lofy KH, Wiesman J, Bruce H, et al. First case of 2019 novel coronavirus in the United States. New England Journal of Medicine. 2020.

8 Ahmadzadeh J, Mobaraki K, Mousavi SJ, Aghazadeh-Attari J, Mirza-Aghazadeh-Attari M, Mohebbi I. The risk factors associated with MERS-CoV patient fatality: A global survey. Diagnostic microbiology and infectious disease. 2020;96(3):114876.

9 Jia N, Feng D, Fang LQ, Richardus JH, Han XN, Cao WC, et al. Case fatality of SARS in mainland China and associated risk factors. Tropical Medicine & International Health. 2009.7–14:21;

10 Team EE. Note from the editors: World Health Organization declares novel coronavirus (2019-nCoV) sixth public health emergency of international concern. Eurosurveillance. 2020;25(5):200131e.

11 Zhao Z, Xie J, Yin M, Yang Y, He H, Jin T, et al. Clinical and Laboratory Profiles of 75 Hospitalized Patients with Novel Coronavirus Disease 2019 in Hefei, China. medRxiv. 2020.

12 Cao M, Zhang D, Wang Y, Lu Y, Zhu X, Li Y, et al. Clinical Features of Patients Infected with the 2019 Novel Coronavirus (COVID-19) in Shanghai, China. medRxiv. 2020.

13 Chen G, Wu D, Guo W, Cao Y, Huang D, Wang H, et al. Clinical and immunologic features in severe and moderate forms of Coronavirus Disease 2019. medRxiv. 2020.

14 Shi H, Han X, Jiang N, Cao Y, Alwalid O, Gu J, et al. Radiological findings from 81 patients with COVID-19 pneumonia in Wuhan, China: a descriptive study. The Lancet Infectious Diseases. 2020.

15 Huang Y, Tu M, Wang S, Chen S, Zhou W, Chen D, et al. Clinical characteristics of laboratory confirmed positive cases of SARS-CoV-2 infection in Wuhan, China: A retrospective single center analysis. Diabetes. 2020;4:11.8.

16 Xu X-W, Wu X-X, Jiang X-G, Xu K-J, Ying L-J, Ma C-L, et al. Clinical findings in a group of patients infected with the 2019 novel coronavirus (SARS-Cov-2) outside of Wuhan, China: retrospective case series. Bmj. 2020;368.

17 Yang W, Cao Q, Qin L, Wang X, Cheng Z, Pan A, et al. Clinical characteristics and imaging manifestations of the 2019 novel coronavirus disease (COVID-19 :(A multi-center study in Wenzhou city, Zhejiang, China. Journal of Infection. 2020.

18 Guan W-j, Ni Z-y, Hu Y, Liang W-h, Ou C-q, He J-x, et al. Clinical characteristics of 2019 novel coronavirus infection in China. MedRxiv. 2020.

19 Ai J, Chen J, Wang Y, Liu X, Fan W, Qu G, et al. The cross-sectional study of hospitalized coronavirus disease 2019 patients in Xiangyang, Hubei province. medRxiv. 2020.

20 Chen X, Zheng F, Qing Y, Ding S, Yang D, Lei C, et al. Epidemiological and clinical features of 291 cases with coronavirus disease 2019 in areas adjacent to Hubei, China: a double-center observational study. medRxiv. 2020.

21 Xu X, Yu C, Qu J, Zhang L, Jiang S, Huang D, et al. Imaging and clinical features of patients with 2019 novel coronavirus SARS-CoV-2. European Journal of Nuclear Medicine and Molecular Imaging. 2020:1–6.

22 Li X, Wang L, Yan S, Yang F, Xiang L, Zhu J, et al. Clinical characteristics of 25 death cases infected with COVID-19 pneumonia: a retrospective review of medical records in a single medical center, Wuhan, China. medRxiv. 2020.

23 Qian G-Q, Yang N-B, Ding F, Ma AHY, Wang Z-Y, Shen Y-F, et al. Epidemiologic and Clinical Characteristics of 91 Hospitalized Patients with COVID-19 in Zhejiang, China: A retrospective, multi-centre case series. medRxiv. 2020.

24 Liang Y, Liang J, Zhou Q, Li X, Lin F, Deng Z, et al. Prevalence and clinical features of 2019 novel coronavirus disease (COVID-19) in the Fever Clinic of a teaching hospital in Beijing: a single-center, retrospective study. medRxiv. 2020.

25 Li J, Zhang Y, Wang F, Liu B, Li H, Tang G, et al. Sex differences in clinical findings among patients with coronavirus disease 2019 (COVID-19) and severe condition. medRxiv. 2020.

26 Huang Y, Zhou H, Yang R, Xu Y, Feng X, Gong P. Clinical characteristics of 36 non-survivors with COVID-19 in Wuhan, China. medRxiv. 2020.

27 Yang P, Ding Y, Xu Z, Pu R, Li P, Yan J, et al. Epidemiological and clinical features of COVID-19 patients with and without pneumonia in Beijing, China. medRxiv. 2020

28 Fu H, Li H, Tang X, Li X, Shen J, Zhou Y, et al. Analysis on the Clinical Characteristics of 36 Cases of Novel Coronavirus Pneumonia in Kunming. medRxiv. 2020.

29 Liu T, Zhang J, Yang Y, Zhang L, Ma H, Li Z, et al. The potential role of IL-6 in monitoring coronavirus disease 2019. Available at SSRN 3548761. 2020.

30 Zhang G, Hu C, Luo L, Fang F, Chen Y, Li J, et al. Clinical features and outcomes of 221 patients with COVID-19 in Wuhan, China. medRxiv. 2020.

31 Xia W, Shao J, Guo Y, Peng X, Li Z, Hu D. Clinical and CT features in pediatric patients with COVID-19 infection: Different points from adults. Pediatric pulmonology. 2020.

32 Wu J, Wu X, Zeng W, Guo D, Fang Z, Chen L, et al. Chest CT findings in patients with corona virus disease 2019 and its relationship with clinical features. Invest Radiol. 2020;10.

33 Wu J, Liu J, Zhao X, Liu C, Wang W, Wang D, et al. Clinical characteristics of imported cases of COVID-19 in Jiangsu province: a multicenter descriptive study. Clinical Infectious Diseases. 2020.

34 Kui L, Fang Y-Y, Deng Y, Liu W, Wang M-F, Ma J-P, et al. Clinical characteristics of novel coronavirus cases in tertiary hospitals in Hubei Province. Chinese medical journal. 2020.

35 Zhou F, Yu T, Du R, Fan G, Liu Y, Liu Z, et al. Clinical course and risk factors for mortality of adult inpatients with COVID-19 in Wuhan, China: a retrospective cohort study. The Lancet. 2020.

36 Zhang F, He L, Ouyang Y, Gong J, Li X, Wei Y, et al. Clinical Features of 81 Hospitalized Patients with 2019 Novel Coronavirus-Infected Pneumonia in Jingzhou, China: A Descriptive Study. 2020.

37 Cai J, Xu J, Lin D, Xu L, Qu Z, Zhang Y, et al. A Case Series of children with 2019 novel coronavirus infection: clinical and epidemiological features. Clinical Infectious Diseases. 2020.

38 Xu Y-H, Dong J-H, An W-M, Lv X-Y, Yin X-P, Zhang J-Z, et al. Clinical and computed tomographic imaging features of Novel Coronavirus Pneumonia caused by SARS-CoV-2. Journal of Infection. 2020.

39 Liu YSL, Zhang D, Tang S, Chen H, Chen L, He X, et al. The Epidemiological and Clinical Characteristics of 2019 Novel Coronavirus Infection in Changsha, China. China (2/10/2020). 2020.

40 Liu W, Wang F, Li G, Wei Y, Li X, He L, et al. Analysis of 2019 Novel Coronavirus Infection and Clinical Characteristics of Outpatients: An Epidemiological Study from the Fever Clinic in Wuhan, China. 2020.

41 Xiong Y, Sun D, Liu Y, Fan Y, Zhao L, Li X, et al. Clinical and High-Resolution CT Features of the Severe Acute Respiratory Syndrome Coronavirus 2 (SARS-CoV-2) Pneumonia: Comparison of the Initial and Follow-Up Changes. Available at SSRN 3539695. 2020.

42 Liu Y, Yang Y, Zhang C, Huang F, Wang F, Yuan J, et al. Clinical and biochemical indexes from 2019-nCoV infected patients linked to viral loads and lung injury. Science China Life Sciences. 2020;63(3):364–74.

43 Lu C, Yang W, Hu Y, Hui J, Zhou G, Shu J, et al. Coronavirus Disease 2019 (COVID-19) Pneumonia: Early Stage Chest CT Imaging Features and Clinical Relevance. 2020.

44 Lei Z-Y, Cao H-J, Jie Y-S, Huang Z-L, Guo X-Y, Chen J-F, et al. Comparison of Epidemiological and Clinical Features of Patients with Coronavirus Disease (COVID-19) in Wuhan and Outside Wuhan, China. 2020.

45 Liu H, Liu F, Li J, Zhang T, Wang D, Lan W. Clinical and CT imaging features of the COVID-19 pneumonia: Focus on pregnant women and children. Journal of infection. 2020.

46 Chen L, Liu H, Liu W, Liu J, Liu K, Shang J, et al. Analysis of clinical features of 29 patients with 2019 novel coronavirus pneumonia. Zhonghua jie he he hu xi za zhi= Zhonghua jiehe he huxi zazhi= Chinese journal of tuberculosis and respiratory diseases. 2020;43:E005–E.

47 Zhu X, Yuan W, Huang K, Wang Q, Yao S, Lu W, et al. Clinical Features and Short-Term Outcomes of 114 Elderly Patients with COVID-19 in Wuhan, China: A Single-Center, Retrospective, Observational Study. China: A Single-Center, Retrospective, Observational Study (3/2/2020). 2020.

48 Li K, Wu J, Wu F, Guo D, Chen L, Fang Z, et al. The clinical and chest CT features associated with severe and critical COVID-19 pneumonia. Investigative radiology. 2020.

49 Wan S, Yi Q, Fan S, Lv J, Zhang X, Guo L, et al. Characteristics of lymphocyte subsets and cytokines in peripheral blood of 123 hospitalized patients with 2019 novel coronavirus pneumonia (NCP). Medrxiv. 2020.

50 Jian-ya G. Clinical characteristics of 51 patients discharged from hospital with COVID-19 in Chongqing, China. medRxiv. 2020.

51 Zhang B, Zhou X, Qiu Y, Feng F, Feng J, Jia Y, et al. Clinical characteristics of 82 death cases with COVID-19. medRxiv. 2020.

52 Cui P, Chen Z, Wang T, Dai J, Zhang J, Ding T, et al. Clinical features and sexual transmission potential of SARS-CoV-2 infected female patients: a descriptive study in Wuhan, China. medRxiv. 2020.

53 Qi D, Yan X, Tang X, Peng J, Yu Q, Feng L, et al. Epidemiological and clinical features of 2019-nCoV acute respiratory disease cases in Chongqing municipality, China: a retrospective, descriptive, multiple-center study. 2020.

54 Xu Y, Xu Z, Liu X, Cai L, Zheng H, Huang Y, et al. Clinical findings in critical ill patients infected with SARS-Cov-2 in Guangdong Province, China: a multi-center, retrospective, observational study. medRxiv. 2020.

55 Liao J, Fan S, Chen J, Wu J, Xu S, Guo Y, et al. Epidemiological and clinical characteristics of COVID-19 in adolescents and young adults. medRxiv. 2020.

56 Lei Y. Clinical features of imported cases of coronavirus disease 2019 in Tibetan patients in the Plateau area. medRxiv. 2020.

57 Zhang C, Gu J, Chen Q, Deng N, Li J, Huang L, et al. Clinical Characteristics of 34 Children with Coronavirus Disease-2019 in the West of China: a Multiple-center Case Series. medRxiv. 2020:2020.03.12.20034686.

58 Liu J, Ouyang L, Guo P, sheng Wu H, Fu P, liang Chen Y, et al. Epidemiological, Clinical Characteristics and Outcome of Medical Staff Infected with COVID-19 in Wuhan, China: A Retrospective Case Series Analysis. medRxiv. 2020.

59 Zhang J-j, Dong X, Cao Y-y, Yuan Y-d, Yang Y-b, Yan Y-q, et al. Clinical characteristics of 140 patients infected with SARS-CoV-2 in Wuhan, China. Allergy. 2020.

60 Pan Y, Ye G, Zeng X, Liu G, Zeng X, Jiang X, et al. Can routine laboratory tests discriminate 2019 novel coronavirus infected pneumonia from other community-acquired pneumonia? medRxiv.2020.

61 Tavan h, Mohammadi i, Carson KV, Sayehmiri K, Sayehmiri f. Prevalence of Epilepsy in Iran: A Meta-Analysis and Systematic Review. Iranian Journal of Child Neurology. 2014;8(4.(

62 Sayehmiri K, Tavan H. Systematic review and meta-analysis methods prevalence of peptic ulcer in IRAN. Journal of Govaresh. 2015;20(4):250–8.

63 Moher D, Shamseer L, Clarke M, Ghersi D, Liberati A, Petticrew M, et al. Preferred reporting items for systematic review and meta-analysis protocols (PRISMA-P) 2015 statement. Systematic reviews. 2015;4(1):1.

64 Luchini C, Stubbs B, Solmi M, Veronese N. Assessing the quality of studies in metaanalyses: Advantages and limitations of the Newcastle Ottawa Scale. World J Meta-Anal. 2017;5(4):80–4.

65 Tarsilla M. Cochrane handbook for systematic reviews of interventions. Journal of Multidisciplinary Evaluation. 2010;6(14):142–8.

66 Ades A, Lu G, Higgins J. The interpretation of random-effects meta-analysis in decision models. Medical Decision Making. 2005;25(6):646–54.

67 Begg CB, Mazumdar M. Operating characteristics of a rank correlation test for publication bias. Biometrics. 1994:1088–101.

68 !!! .INVALID CITATION!!!

69 Rodriguez-Morales AJ, Cardona-Ospina JA, Gutiérrez-Ocampo E, Villamizar-Peña r, Holguin-Rivera Y, Escalera-Antezana JP, et al. Clinical, laboratory and imaging features of COVID-19: A systematic review and meta-analysis. Travel medicine and infectious disease. 2020:101623.

70 Cao Y, Liu X, Xiong L, Cai K. Imaging and clinical features of patients with 2019 novel coronavirus SARS-CoV-2: a systematic review and meta-analysis. Journal of medical virology. 2020.

71 Leung WK, To K-f, Chan PK, Chan HL, Wu AK, Lee N, et al. Enteric involvement of severe acute respiratory syndrome-associated coronavirus infection. Gastroenterology. 2003;125(4):1011–7.

72 Peiris JSM, Chu C-M, Cheng VC-C, Chan K, Hung I, Poon LL, et al. Clinical progression and viral load in a community outbreak of coronavirus-associated SARS pneumonia: a prospective study. The Lancet. 2003;3.72-1767:(9371)61

73 Assiri A, McGeer A, Perl TM, Price CS, Al Rabeeah AA, Cummings DA, et al. Hospital outbreak of Middle East respiratory syndrome coronavirus. New England Journal of Medicine. 2013;369(5):407–16.

74 Xu Z, Shi L, Wang Y, Zhang J, Huang L, Zhang C, et al. Pathological findings of COVID-19 associated with acute respiratory distress syndrome. The Lancet respiratory medicine. 2020;8(4):420–2.

75 Zhou P, Yang X-L, Wang X-G, Hu B, Zhang L, Zhang W, et al. A pneumonia outbreak associated with a new coronavirus of probable bat origin. nature. 2020;579(7798):270–3.

76 Xu X, Gao X-M. Immunological responses against SARS-coronavirus infection in humans. Cell Mol Immunol. 2004;1(2):119–22.

77 Frasca L, Piazza C, Piccolella E. CD4+ T cells orchestrate both amplification and deletion of CD8+ T cells. Critical Reviews™ in Immunology. 1998;18(6.(

78 Ng CT, Snell LM, Brooks DG, Oldstone MB. Networking at the level of host immunity: immune cell interactions during persistent viral infections. Cell host & microbe. 2013;13(6):652– 64.

79 Fenwick C, Joo V, Jacquier P, Noto A, Banga R, Perreau M, et al. T-cell exhaustion in HIV infection. Immunological Reviews. 2019;292(1):149–63.

80 Diao B, Wang C, Tan Y, Chen X, Liu Y, Ning L, et al. Reduction and functional exhaustion of T cells in patients with coronavirus disease 2019 (COVID-19). Frontiers in Immunology. 2020;11:827.

81 Bonig H, Papayannopoulou T. Mobilization of hematopoietic stem/progenitor cells: general principles and molecular mechanisms. Stem Cell Mobilization: Springer; 2012. p. 1–14.

82 Pepys MB, Hirschfield GM. C-reactive protein: a critical update. The Journal of clinical investigation. 2003;111(12):1805–12.

83 Hahn W-H, Song J-H, Kim H, Park S. Is procalcitonin to C-reactive protein ratio useful for the detection of late onset neonatal sepsis? The Journal of Maternal-Fetal & Neonatal Medicine. 2018;31(6):822–6.

84 Ling W. C-reactive protein levels in the early stage of COVID-19. Medecine et Maladies Infectieuses. 2020.

85 Tan C, Huang Y, Shi F, Tan K, Ma Q, Chen Y, et al. C-reactive protein correlates with CT findings and predicts severe COVID-19 early. Journal of Medical Virology. 2020.

86 Saeed K, Dale A, Leung E, Cusack T, Mohamed F, Lockyer G, et al. Procalcitonin levels predict infectious complications and response to treatment in patients undergoing cytoreductive surgery for peritoneal malignancy. European Journal of Surgical Oncology (EJSO). 2016;42(2):234–43.

87 Choi JJ, McCarthy MW. Novel applications for serum procalcitonin testing in clinical practice. Expert review of molecular diagnostics. 2018;18(1):27–34.

88 Zhang L, Zhang X. Serum sTREM-1, PCT, CRP, Lac as biomarkers for death risk within 28 days in patients with severe sepsis. Open life sciences. 2018;13(1):42.7-

89 Rodríguez A, Reyes L, Monclou J, Suberviola B, Bodí M, Sirgo G, et al. Relationship between acute kidney injury and serum procalcitonin (PCT) concentration in critically ill patients with influenza infection. Medicina intensiva. 2018;42(7):399–408.

90 Paniagua M, Cartmel D, Dominguez C. Receptor Recognition by the Novel Coronavirus from Wuhan: an Analysis Based on Decade-Long Structural Studies of SARS. 2020.

91 Guan G, Gao L, Wang J, Wen X, Mao T, Peng S, et al. Exploring the mechanism of liver enzyme abnormalities in patients with novel coronavirus-infected pneumonia. Zhonghua gan zang bing za zhi= Zhonghua ganzangbing zazhi= Chinese journal of hepatology. 2020;28(2):E002–E.

92 Li F. Structure, function, and evolution of coronavirus spike proteins. Annual review of virology. 2016;3:237–61.

93 Bosch BJ, van der Zee R, de Haan CA, Rottier PJ. The coronavirus spike protein is a class I virus fusion protein: structural and functional characterization of the fusion core complex. Journal of virology. 2003;77(16):8801–11.

94 Matthay MA, Ware LB, Zimmerman GA. The acute respiratory distress syndrome. The Journal of clinical investigation. 2012;122(8):2731–40.

95 Channappanavar R, Perlman S, editors. Pathogenic human coronavirus infections: causes and consequences of cytokine storm and immunopathology. Seminars in immunopathology; 2017: Springer.

96 Huang KJ, Su IJ, Theron M, Wu YC, Lai SK, Liu CC, et al. An interferon-γ-related cytokine storm in SARS patients. Journal of medical virology. 2005;75.94–185:(2)

97 Lippi G, Lavie CJ, Sanchis-Gomar F. Cardiac troponin I in patients with coronavirus disease 2019 (COVID-19): Evidence from a meta-analysis. Progress in cardiovascular diseases. 2020.

98 Li W, Moore MJ, Vasilieva N, Sui J, Wong SK, Berne MA, et al. Angiotensin-converting enzyme 2 is a functional receptor for the SARS coronavirus. Nature. 2003;426(6965):450–4.

99 Cheng Y, Luo R, Wang K, Zhang M, Wang Z, Dong L, et al. Kidney impairment is associated with in-hospital death of COVID-19 patients. MedRxiv. 2020.

100 Henry BM, de Oliveira Mhs, Benoit J, Lippi G. Gastrointestinal symptoms associated with severity of coronavirus disease 2019 (COVID-19): a pooled analysis. Internal and Emergency Medicine. 2020:1.

101 Minasyan H, Flachsbart F. Blood coagulation: a powerful bactericidal mechanism of human innate immunity. International reviews of immunology. 2019;38(1):3–17.

102 Gershom E, Sutherland M, Lollar P, Pryzdial E. Involvement of the contact phase and intrinsic pathway in herpes simplex virus-initiated plasma coagulation. Journal of Thrombosis and Haemostasis. 2010;8(5):1037–43.

103 Rapala-Kozik M, Karkowska J, Jacher A, Golda A, Barbasz A, Guevara-Lora I, et al. Kininogen adsorption to the cell surface of Candida spp. International immunopharmacology. 2008;8(2):237–41.

104 Loof TG, Mörgelin M, Johansson L, Oehmcke S, Olin AI, Dickneite G, et al. Coagulation, an ancestral serine protease cascade, exerts a novel function in early immune defense. Blood, The Journal of the American Society of Hematology. 2011;118(9):2589–98.

105 Kawano N, Wada H, Uchiyama T, Kawasugi K, Madoiwa S, Takezako N, et al. Analysis of the association between resolution of disseminated intravascular coagulation (DIC) and treatment outcomes in post-marketing surveillance of thrombomodulin alpha for DIC with infectious disease and with hematological malignancy by organ failure. Thrombosis Journal. 2020;18(1):2.

106 Han H, Yang L, Liu R, Liu F, Wu K-l, Li J, et al. Prominent changes in blood coagulation of patients with SARS-CoV-2 infection. Clinical Chemistry and Laboratory Medicine (CCLM). 2020;1(ahead-of-print.(

